# Inferring vaccine efficacy and mode of action from human challenge studies

**DOI:** 10.1101/2023.09.24.23296054

**Authors:** Fuminari Miura, Don Klinkenberg, Kylie E.C. Ainslie, Jantien A. Backer, Ka Yin Leung, Scott A. McDonald, Pieter de Boer, Jacco Wallinga

## Abstract

To assess the long-term effect of a vaccination programme, understanding both the efficacy and the mode of action of the vaccine is crucial. The actual mode of action is difficult to infer from field trials, because of the heterogeneity of exposure to infection. Here we show an approach to determine both vaccine efficacy and the mode of action of vaccines from human challenge studies. Our approach reveals how vaccines alter an individual’s susceptibility, and identifies a mixture of different modes of action as a function of the challenged dose. By applying the proposed method to influenza challenge data, we illustrate that potentially the lowest vaccine efficacy may occur at intermediate doses, suggesting a hypothesis that moderate transmission intensities might be optimal for establishing infections in vaccinated individuals.

**One sentence summary:** Human challenge studies reveal the protection mechanism of vaccines by measuring variation in susceptibility to infection.

## Introduction

Emerging infectious diseases have posed great threats to public health for decades, and the importance of decision making based on empirical evidence has been repeatedly shown during emergencies such as the recent pandemic of influenza A (H1N1) in 2009 ^1^ and the current pandemic of COVID-19 ^2^. Although the spread of an epidemic can be partially controlled by non-pharmaceutical interventions (e.g., contact tracing, quarantine, travel restrictions, mask wearing) ^3^, effective mitigation strategies are needed to protect vulnerable people such as the elderly in the long term, given societal and economic constraints. To this end, vaccination is the most promising strategy for sustainably suppressing epidemics. Vaccination programmes for COVID-19 were a successful example of showing the effectiveness of speedy development of vaccines and their early implementations ^4^.

While (short term) vaccine efficacy – measured as the proportional reduction in infection risk of vaccinated *versus* unvaccinated individuals during the course of a clinical trial – attracts much attention, the mode of action of vaccines is crucial to assess the longer term impact of a vaccination campaign on transmission dynamics ^5^. The mode of action is conceptualized by two extremes: one extreme being perfect protection from infection for a proportion of the vaccinated persons while the other proportion is left as susceptible as unvaccinated persons (“all-or-nothing”), and the other extreme being that each vaccinated person has the same reduced probability of infection during each exposure event (“leaky”) ^6,7^. In terms of biological interpretations, the all-or-nothing protection may occur due to the inability of a human host to respond to vaccination (i.e., primary vaccine failure), and the leaky protection may be determined by the degree of immune activation or the match between the vaccine and target pathogens ^8^. These two extremes make clear that the chance of acquiring infection will keep rising with an increasing exposure to infection if the vaccine’s mode of action is “leaky”, but not if the vaccine is “all-or-nothing”. Given the same vaccine efficacy as measured in a field trial, the effectiveness of a vaccination campaign with a “leaky” vaccine would be less than with an “all-or-nothing” vaccine. The actual mode of action might be in between these extremes. While the mode of action of vaccines for various diseases such as influenza ^9^, Malaria ^10^, and HIV in non-human primates ^11^ has been explored based on theoretical arguments, inference of the mode of action in randomized controlled vaccine trials or in observational studies is challenging, as it is difficult to measure the exact (cumulative) dose of the infectious agent to which an individual is exposed.

The exposure to infection can be controlled in human challenge trials where healthy volunteers are exposed to a known challenge dose of the pathogen, and this has potential to be an informative study design for both vaccine efficacy and mode of action of the vaccine. Human challenge trials have been conducted for more than two centuries and are often used for evaluating the efficacies of therapies or vaccines, measured as a reduction in infection risk (i.e., the proportion of infected participants among those challenged by a single dose) between control and treatment arms ^12^. They allow for studying the relationship between challenge dose and infection risk ^13^, and such analysis can elucidate individual heterogeneity in susceptibility to infection and variation in conferred efficacy of vaccines ^14,15^. The difference in individual levels of susceptibility in the study population can be described by probability distributions. These can have various shapes; ranging from sharply peaked to uniform ^14,15^ or even including two peaks ^16^. A flexible description of various shapes of susceptibility is crucial to appropriately understand how vaccines induce their protection mechanisms.

Here we propose an approach to determine both vaccine efficacy and the mode of action of vaccines from observed frequencies of infection of healthy and vaccinated volunteers challenged to various doses of virus. As a motivating example, we illustrate this approach with challenge data from studies evaluating influenza viruses with vaccinated and unvaccinated human volunteers, since influenza has the richest trial data available for both arms.

## Results

### Vaccine efficacy

In our approach, we measure the effect of vaccines as the change in biological susceptibility to infection. We allow for individuals to have different levels of susceptibility, and we refer to an individual’s level as *x* and the distribution of these levels over the population of unvaccinated individuals as *f*(*x*). The interpretation of a level of susceptibility is that an individual with susceptibility level *x = x′* has an *x′* times higher probability of infection given exposure to a single pathogen, compared to an individual that has level *x =* 1. We can mathematically show that, under mild assumptions, the distribution of susceptibility *f*(*x*) determines the probability of escaping infection *ℱ*(*d*) as a function of dose *d*:

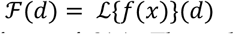

where *ℒ*{*f*(*x*)} is the Laplace transform of *f*(*x*). Thus, the probability of infection among unvaccinated individuals (i.e., the control arm) is the complement of the probability of escaping infection, *P*_*con*_(*d*) *=* 1 − *ℱ*(*d*). By fitting and comparing a set of alternative dose-response curves to the frequencies of infection at various doses as observed in the challenge study, we can infer the distribution of susceptibility levels in the population (**Fig-1A**).

**Fig 1.**
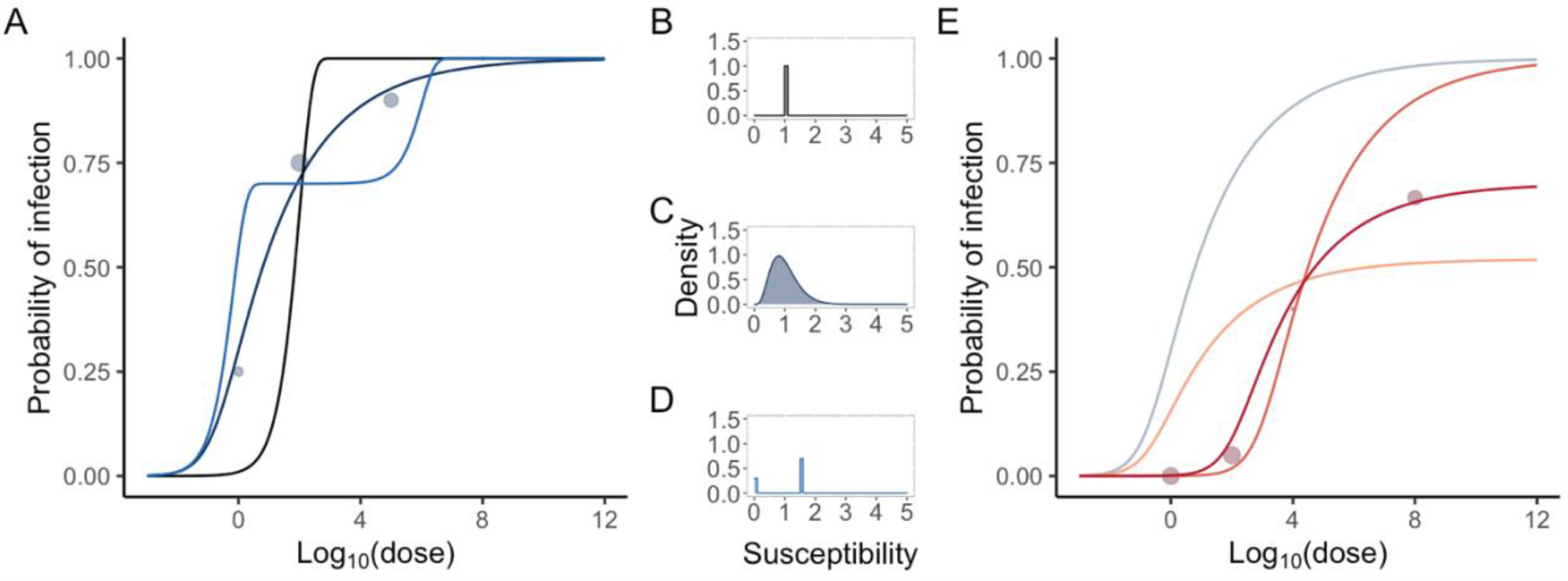
Schematic illustration of the framework for inferring susceptibility distributions via dose-response analysis. In panel (A), bubble plots are the observed human challenge data, and colored lines depict the fitted dose-response curves (colors matching the distributions in panels (B)-(D)). As a result of model fits, we can infer how the susceptibility of an individual is distributed. Panel (B) indicates the case where all individuals have the same level of susceptibility, while Panel (C) shows the case where there is heterogeneity in susceptibility, which is distributed as a Gamma distribution. If a certain proportion is completely immune and the other proportion has one level of susceptibility, a discrete distribution, such as the one in Panel (D), is the most appropriate distribution. Panel (E) illustrates how vaccination changes the dose-response curve, from an unvaccinated state (blue) to vaccinated states (red ones). If the mode of action is “all-or-nothing”, the curve is shrunken along the y-axis (from blue to orange), and if “leaky”, the curve is expanded along the x-axis (blue to light red). The actual effect might be between (“mixture”, shown in dark red). Note that these figures are not actual results of model fits but just a visualization of the concept of this study.

In the same manner, we can describe the probability of escaping infection amongst vaccinated volunteers *ℒ*{*g*(*x*)}(*d*) *= 𝒢*(*d*), for any challenge dose *d*. The probability of infection for vaccinated individuals is then *P*_*vac*_(*d*) *=* 1 − *𝒢*(*d*) where *g*(*x*) is the distribution of susceptibility of vaccinated individuals.

Here we can define the dose-dependent vaccine efficacy against infection in the usual way:

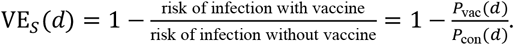

### Mode of action of vaccines

#### (i) All-or-nothing

If the vaccine elicits perfect protection from infection with probability *ϕ*_*v*_, and no protection otherwise, the distribution of susceptibility levels of vaccinated persons *g*(*x*) becomes a mixture of two distributions, one for those who are perfectly protected and one for those who are not protected at all, *g*_*AoN*_(*x*) *= ϕ*_*v*_*δ*(*x*) + (1 − *ϕ*_*v*_)*f*(*x*) where *δ*(*x*) represents the Dirac delta function. The probability of infection for vaccinated individuals at a given dose *P*_*vac*_(*d*) can be obtained from the probability of infection for unvaccinated individuals *P*_*con*_(*d*) by shrinking the probability of infection towards zero by a factor 1 − *ϕ*_*v*_ (Fig 1B):

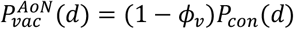

#### (ii) Leaky

If the vaccine acts as a leaky vaccine (i.e., a mode of vaccine that reduces the probability of infection given exposure by a factor of relative reduction *ψ*_*v*_, for each vaccinated person), *P*_vac_(*d*) is expressed as a function which expands *P*_con_(*d*) along the x-axis (for dose):

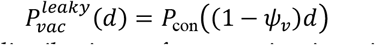

where the susceptibility distribution after vaccination is written as 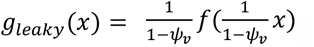.

#### (iii) Mixture of all-or-nothing and leaky

If a vaccine acts as a combination of all-or-nothing and leaky, a fraction *ϕ*_*v*_ is perfectly protected and the other fraction 1 − *ϕ*_*v*_ is partially protected by the leaky effect, resulting in the reduced dose (1 − *ϕ*_*v*_)*d*. The probability of infection for a vaccinated individual is written as:

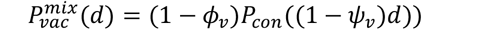

where the susceptibility distribution becomes 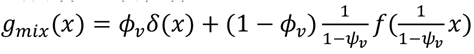.

#### (iv) The generalized form

Above mechanisms (i)-(iii) describe the distribution of susceptibility among the vaccinated, *g*(*x*), related to that of unvaccinated individuals. The assumption here is that the background susceptibility *f*(*x*) is preserved and that *g*(*x*) is expressed by changing *f*(*x*) with factors *ϕ*_*v*_ and *ψ*_*v*_. If it is likely that the vaccine changes the shape of distributions of background susceptibility completely, we need to estimate *g*(*x*) directly from the observations:

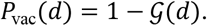

If there is sufficient observed data, we can examine any type of distribution with this generalized form. As a generalized model, the Tweedie model is a natural candidate for describing continuous and positive data with an arbitrary measure scale; it is an exponential dispersion model which is closed under re-scaling ^17^. In addition, it is especially appealing because modelling continuous variables with exact zeros is possible and it has a simple analytic Laplace transform. Thus, we propose a susceptibility distribution *x ∼* Tweedie(*λ, α,γ*), as a generalized model. The parameter *λ* is a direct estimator for infection-blocking immunity, and therefore the all-or-nothing effect in models (i) and (iii) (namely, *ϕ*_*v*_ *=* 1 − *exp*(−*λ*)).

### Vaccine efficacy and mode of action of influenza vaccines as inferred from challenge data

We collected datasets from published studies, with the numbers of infected and uninfected participants after challenge with a known dose of influenza virus. These challenge trials differed in strains, subtypes and types of influenza virus, and in challenge procedures. **Table S3** and **Table S4** provide a summary of the full dataset for unvaccinated and vaccinated individuals, respectively. As a result of literature review, 14 studies were found, and the complete data of all trials were used for fitting of dose-response curves.

The distribution of susceptibility levels for unvaccinated individuals was best described by a gamma distribution, in terms of the Akaike information criterion (AIC), and the model comparison is shown in **Table S1**, and **Table S2**. The distribution of susceptibility levels of vaccinated individuals was best described by the generalized model (iv) (i.e., a Tweedie model)(**Fig-2**). The proportion of individuals who are perfectly protected by vaccine (1 − *exp*(−*λ*)) was estimated to be 0.48 [95%CI: 0.42,0.56]; the second best fitted model was as an all-or-nothing vaccine with an efficacy of 0.48 [95%CI: 0.41,0.55] (**Table-1**). Both suggest that the mode of action of influenza vaccines for influenza virus may be “all-or-nothing”. Note that the estimated confidence intervals for parameters of leaky effects in these models (i.e., *ψ*_*v*_ in model (iii) and (*α,γ*) in model (iv)) were wide, suggesting that the observed data contains limited information on the leaky effect (**Table-1**).

**Fig 2.**
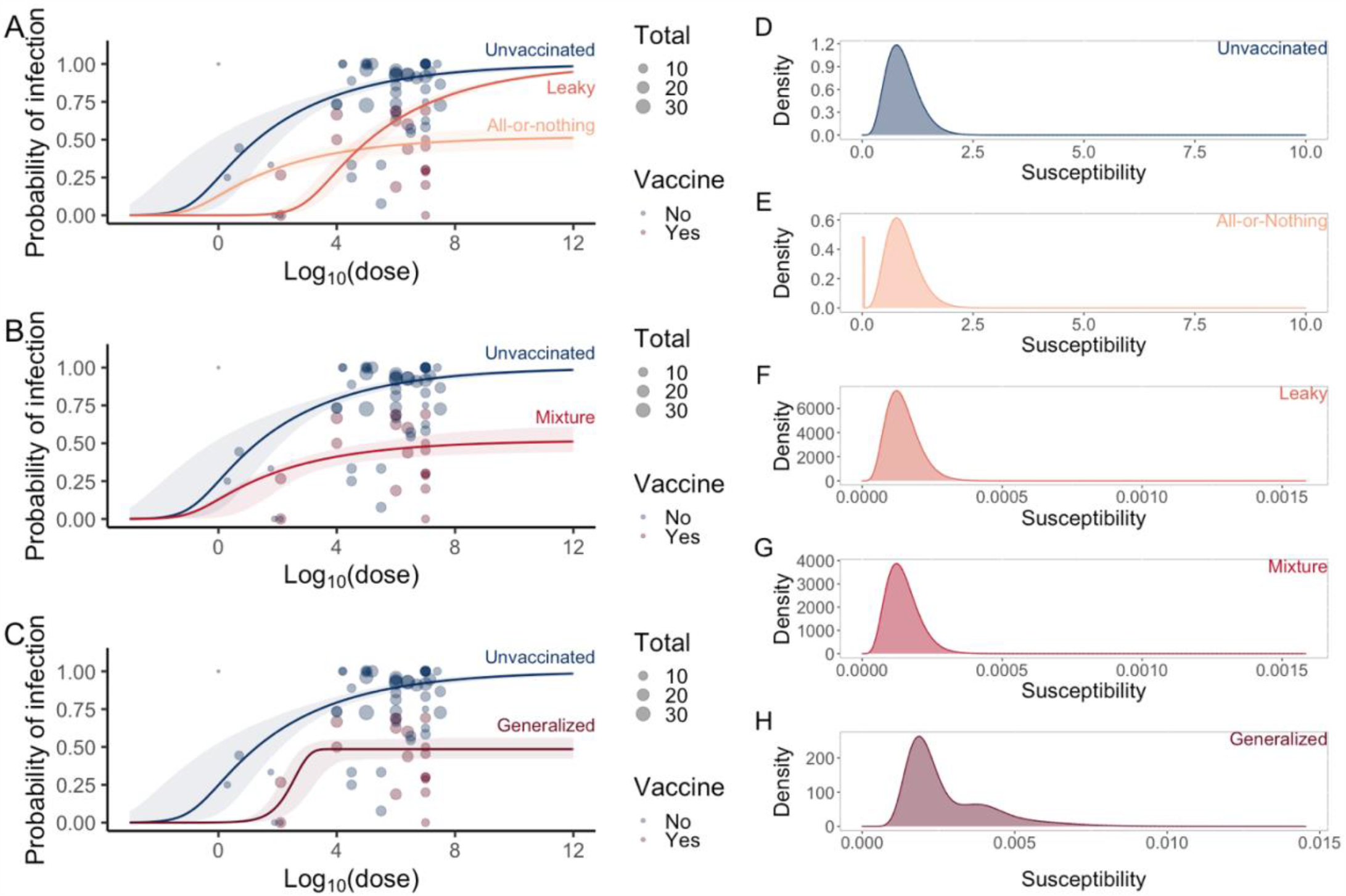
Estimation of dose-response curves for unvaccinated (control) individuals and individuals vaccinated by all-or-nothing and leaky vaccines. Bubble plots are observed challenge data, and the size of bubbles indicates the number of participants in each trial. Panel (A) shows the fitted models of unvaccinated (control), all-or-nothing (model(i)), leaky (model(ii)), and mixture (model (iii)). Panel (B) shows the generalized model of vaccinated individuals (model (iv), Tweedie model) compared to the unvaccinated model. Panels (C)-(G) illustrate the estimated distributions of susceptibility in each model. The reduction in susceptibility due to vaccination is shown in two ways in the distributions; the fraction who acquired all-or-nothing protection is expressed as the point mass at zero (e.g., (D)), and the leaky effect is shown as scaling of both axes (e.g., (E)). In the case of (E), the estimated parameter, 1 − *ψ*_*v*_ *=* 1.6 × 10^−4^, is the scaling factor for both axes.

**Table 1.** Model performances with challenge data of vaccinated individuals. Values in square brackets indicate 95% bootstrapped confidence intervals.

|  | Control | Model (i) | Model (ii) | Model (iii) | Model (iv) |
| --- | --- | --- | --- | --- | --- |
| Mode of action | Unvaccinated | All-or-nothing | Leaky | Mix | Generalized |
| Background distribution $f(x)$ | $x \sim \text{Gamma}(k, \theta)$ | $x \sim \text{Gamma}(k, \theta)$ | $x \sim \text{Gamma}(k, \theta)$ | $x \sim \text{Gamma}(k, \theta)$ | Not applicable |
| Vaccinated distribution $g(x)$ | Not applicable | $\phi_v \delta(x) + (1 - \phi_v) f(x)$ | $\frac{1}{1 - \psi_v} f(\frac{1}{1 - \psi_v} x)$ | $\phi_v \delta(x) + \frac{1 - \phi_v}{1 - \psi_v} f(\frac{1}{1 - \psi_v} x)$ | $x \sim \text{Tweedie}(\lambda, \alpha, \gamma)$ |
| Estimated parameters | - | $\phi_v = 0.48$<br>[95%CI: 0.41,0.55] | $1 - \psi_v = 1.6 \times 10^{-4}$<br>[95%CI: $5.4 \times 10^{-5}$, $4.6 \times 10^{-4}$] | $\phi_v = 0.48$<br>[95%CI: 0.38, 0.55]<br>$1 - \psi_v = 1$<br>[95%CI: $1.7 \times 10^{-2}$, 1] | $\lambda = 0.66$<br>[95%CI: 0.55,0.83]<br>$\alpha = 1.4 \times 10^{-4}$<br>[95%CI: $1.5 \times 10^{-8}$, $2.8 \times 10^{-3}$]<br>$\gamma = 14$<br>[95%CI: $3.2 \times 10^{-1}$, $2.3 \times 10^4$] |
| Log-likelihood | - | -159.9 | -187.9 | -159.9 | -155.7 |
| AIC | - | 321.9 | 377.8 | 323.9 | 317.4 |

The estimated vaccine efficacy for “all-or-nothing” vaccines is constant over dose because of its perfect protection mechanism (**Fig-3A**). By contrast, the efficacy of “leaky” vaccines decreases as the dose increases; the reason is that the probability of infection of both control and vaccinated groups will converge to the same value when the dose is high enough. As a result, an all-or-nothing effect can best be estimated from trials with many high challenge doses, whereas a good estimate of the leaky effect requires trial data with low inoculation doses. Because the published studies were mainly done with high doses, the parameters for the leaky effect were not identifiable (wide confidence intervals). Interestingly, the generalized model that best describes the data suggests a vaccine efficacy that first decreases to an estimated VE_*S*_(*d*) of 0.39 at the dose of 10^4.0^ (**Fig-3D**), and then increases.

**Fig 3.**
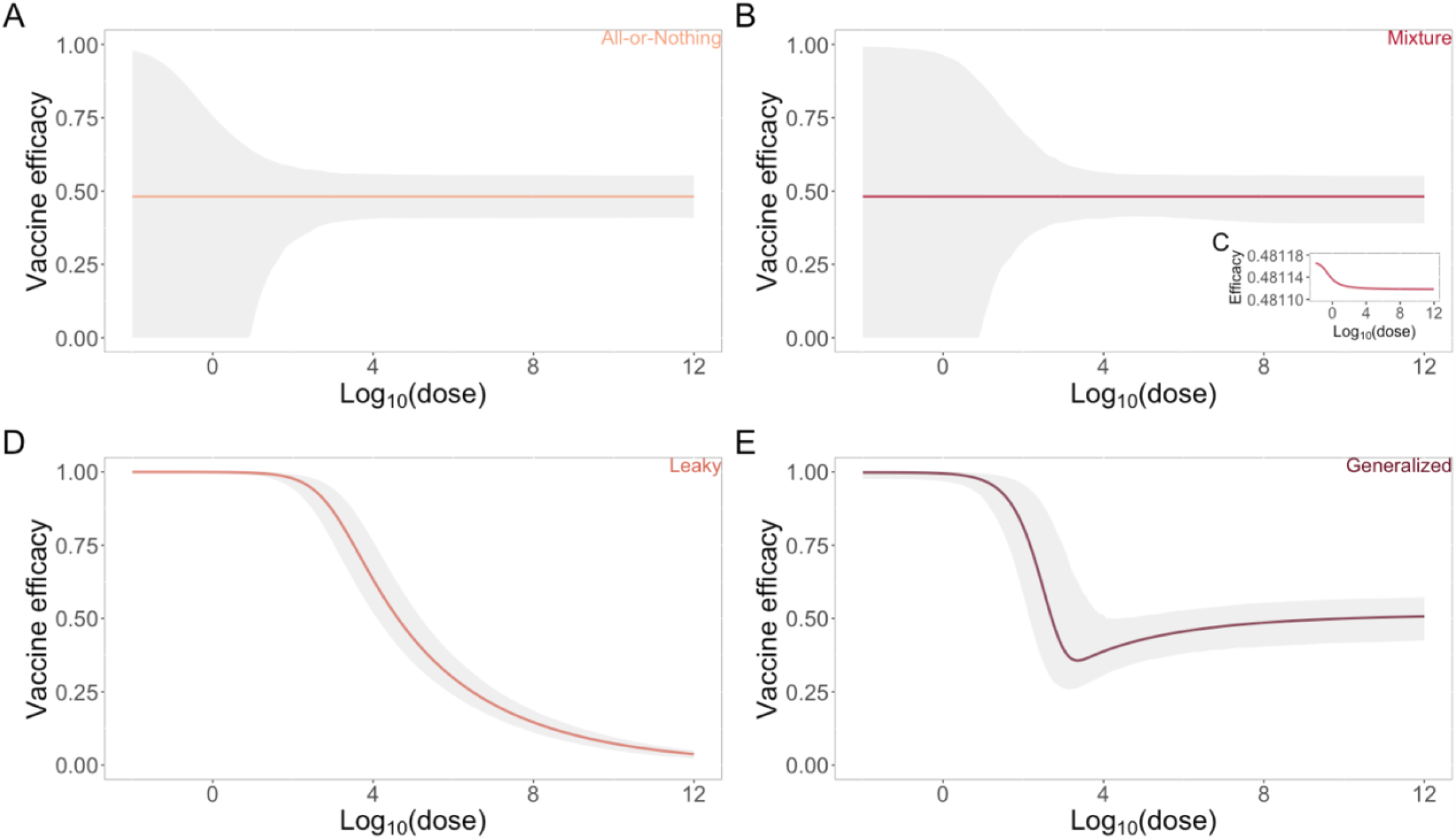
Estimated dose-dependent vaccine efficacy. Relative risk (RR) is defined as the ratio of the probability of infection with vaccination to the probability of infection without vaccination, and the vaccine efficacy is defined as 1 - RR. Gray areas indicate 95% bootstrapped confidence intervals. To draw the uncertainty, we generated 1000 bootstrap samples of parameters of both unvaccinated and vaccinated dose-response models, and took the (dose-dependent) risk ratio. Each panel shows the estimated vaccine efficacy for four different models; all-or-nothing (A), mixture ((B) and (C)), leaky (D), and generalized (E)models.

## Discussion

In this study, we showed how to infer both the vaccine efficacy and the mode of action from human challenge trials on vaccinated and unvaccinated individuals. With the proposed method, we measured the effect of vaccines as the change in biological susceptibility to infection, from a relatively small number of observed infections.

Human challenge studies have an advantage over the standard vaccine field trials, as it is easier to control for unmeasured covariates, and the infection process (i.e., exposure, infection, illness onset, and virus shedding) can be directly observed. A crucial advantage of challenge trials is that there is no unobservable heterogeneous exposure to infection, which causes bias in the estimate of VE ^8,18^. With controlled exposures in challenge trials, our proposed method can measure the heterogeneity in susceptibility.

Measuring the vaccine efficacy as the change in susceptibility distribution answers to the general question about how to infer the mode of action. Especially in the context of COVID-19, empirical evidence on the mode of action of vaccines is crucial for the optimization of vaccine allocations with current modelling schemes ^19,20^. Although one challenge trial for SARS-CoV-2 became public ^21^, there are still ongoing registrations of challenge trials over the world ^22,23^. Our method can be easily implemented in those trials. To maximize the benefit of those studies, our results implied two important suggestions.

First, the low dose region is essential to estimate the distribution of susceptibility levels and to infer vaccine efficacy and mode of action, although many trials tend to use high doses. Dose-response analysis requires multiple data points that cover a wide range of doses, and our analysis with influenza data also showed that models failed to converge or became stuck in a local minimum when low dose data are not available. These results indicated that low dose data increases the (practical) identifiability of model parameters. Second, trials with volunteers who have been vaccinated are also informative. Several vaccines against COVID-19 have been introduced into the population at different speeds, and thus it will be difficult to set up well-controlled field trials ^24^. Even in such cases, the proposed method can provide supportive evidence to judge the efficacy and the safety of vaccines.

Vaccine efficacy is not necessarily always dose-independent (all-or-nothing), nor does it need to monotonically decline with dose (leaky). Use of the more general Tweedie model does create room for alternatives, such as a minimum vaccine efficacy for intermediate exposure doses, as Figure 3D would suggest. However, the existence of such a minimum for influenza cannot be established with the currently available data, as the leaky effect could not reliably be estimated. The estimated range of VEs from 0.39 to 0.51 is consistent to the results of previous randomized controlled trials; for example, a meta-analysis showed that the pooled efficacy was 59 % based on published data from 1967 to 2011 ^25^. If the exposed dose per contact in the natural infection setting is measurable, the dose-dependent VE estimated by this method would be comparable to vaccine effectiveness observed in field trials.

There are several drawbacks in this scheme. First, as pointed out in the literature ^26^, inoculation methods (intramuscular, aerosol, etc.) do not always perfectly represent natural infections, and thus the protection efficacy might differ in actual exposure settings. Although we apply the proposed method to the pooled data that include different influenza strains, stratified analysis is needed when such data become available. Second, the study population is biased because participants who enroll in challenge trials are mainly healthy adults. When we generalize the obtained results to other populations, such as subgroups at high risk from infection, the extrapolation should be carefully done, combining with other supportive observations ^16^. Third, our method assumes that any single pathogen particle is independent and can establish infection (single hit theory ^13^). While this assumption is considered to be reasonable in general, further investigations such as cell-based infection experiments are needed. Alternatively, challenge trials could be designed with series of smaller doses compared to single large doses. Yet, we argue that challenge trials are the most well-controlled design to measure the efficacy and the mode of actions of vaccines, and thus can provide useful information for long-term effects of vaccination campaigns, especially when combined with field trials measuring short-term effectiveness in the population.

In conclusion, we have described the principle for measuring vaccine efficacy as the change in the distribution of susceptibility and how to specify the mode of action of vaccines with human challenge data. The presented results emphasized the importance of data sharing for outcomes across multiple trials, such as upcoming COVID-19 trials ^27^. The open science movement would maximize the benefit of those trials and decipher the epidemiology of emerging diseases.

## Data Availability

All codes and analyzed data are available at the authors' GitHub link (https://github.com/fmiura/VacMoA_2023).

## Acknowledgements

We thank Susan van den Hof for reviewing the manuscript.

## Author contributions

Conceptualization: FM, JW.

Data curation: FM.

Formal analysis: FM, JW.

Investigation: FM.

Methodology: FM, DK, JW.

Software: FM.

Validation: FM, DK, JW.

Visualization: FM.

Writing – original draft: FM, DK, KECA, JAB, KYL, SAM, PB, JW.

Writing – review & editing: FM, DK, KECA, JAB, KYL, SAM, PB, JW.

## Materials and Methods

### Human challenge data

Here we show the challenge data of influenza viruses collected from the published articles. The data used in our further analysis are composed of the number of exposed doses, total participants, and infected individuals in each trial. Infection status was determined by antibody level or the presence of viruses. **Table S3** and **Table S4** provide a summary of the analyzed dataset. It should be noted that reported strains are different among studies, and the condition of inoculated samples might differ. Several previous studies argued that the aggregation of virus particles might affect the result of dose-response analysis ^13,26^. Therefore, when human study data are synthesized, we need to look through the experimental process written in the original articles and determine the unit of dose, depending on the treatment process of inoculation samples. The availability of those details is also summarized in **Table S3** and **Table S4**.

### Dose-response analysis

Here we denote the probability of infection in controlled infection experiments as

*P*(*d*), a function of dose *d*. Given that the susceptibility of unvaccinated individuals differs and is distributed as *f*(*x*) with a level of susceptibility *x*, the probability of infection by a dose *d* for an (control) individual is written as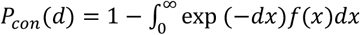. The interpretation of variable *x* is that an individual with the level of susceptibility *x = x′* has *x′* times higher probability of infection compared to an individual with *x =* 1. To estimate a set of unknown parameters ***θ*** of *f*(*x*), the binomial likelihood of all experiment groups is

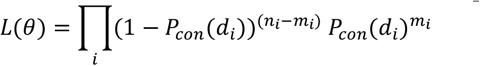

and thus the log-likelihood is

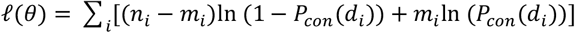

where for each trial *i* we have a dose *d*_*i*_ and a group of *n*_*i*_ volunteers of which *m*_*i*_ are infected. We assumed all the particles are infectious and effective to establish an infection (single-hit theory ^13^). The above likelihood was used for the parameter inference of susceptibility distributions among both unvaccinated and vaccinated individuals (i.e., *f*(*x*) and *g*(*x*), respectively). We performed the maximum likelihood estimation (MLE) using the optim() function in the R statistical programming environment version 3.5.1. ^28^, and 95 % confidence intervals were computed from the bootstrapped samples.

### Derivation of the susceptibility distributions of vaccinated individuals *g*(*x*)

The susceptibility distribution of vaccinated individuals *g*(*x*) is written as the change from that of unvaccinated, such as

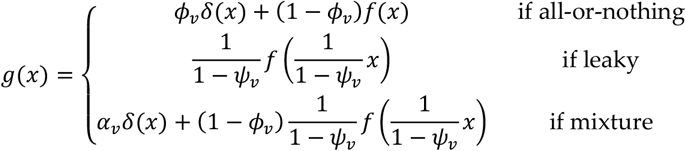

These are analytically derived, through the scaling law and linearity property of Laplace transform.

*Proof*.

i. All-or-nothing

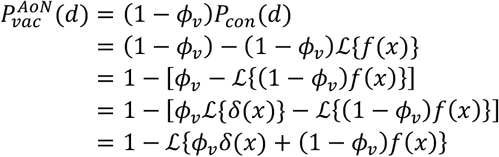
ii. Leaky

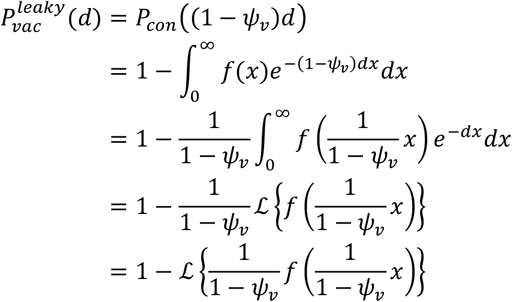
iii. Mixture

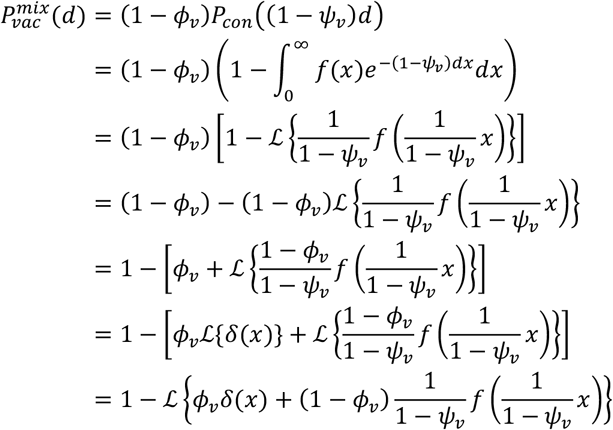

If there is sufficient observed data, we can directly estimate the susceptibility distribution for vaccinated individuals, *g*(*x*), by allowing *g*(*x*) to be any distribution. We can simply write the generalized dose-response model as

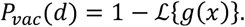

While any distribution is applicable, as a generalized model, a Tweedie distribution is a natural candidate for describing continuous and positive data with an arbitrary measure scale. A Tweedie model, which is a compound Poisson Gamma model, can have exact zeros and shifts from a Poisson distribution to a gamma distribution as the variance power 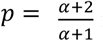 moves from 1 to 2. When *p =* 1, it becomes a Poisson distribution. When *p =* 2, it becomes a gamma distribution. A Tweedie model is useful to seek for the plausible mixture effect of vaccines, considering both leaky and all-or-nothing effects. The probability of infection of vaccinated individuals is written as a complement of the Laplace transform;

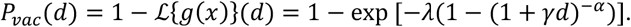

### Other examined distributions

To capture the heterogeneity in susceptibility among unvaccinated individuals, there are several distributions that explain different biological background mechanisms. We examine six different models to estimate the susceptibility distribution for unvaccinated individuals *f*(*x*), and details of those models are below:

#### One-level (Delta) model

If all individuals have the same level of susceptibility, we can express it as:

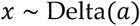

and, using its Laplace transform, the probability of infection is

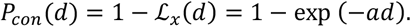

#### Two-level model

If a population has two levels of susceptibility, a proportion *p*_1_ has the level *a*_1_ while the other proportion (1 − *p*_1_) has the level *a*_2_:

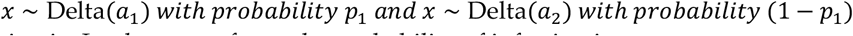

and, using its Laplace transform, the probability of infection is

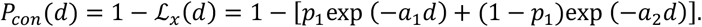

#### Gamma model

If the level of susceptibility for all individuals follows gamma distribution:

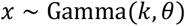

and, using its Laplace transform, the probability of infection is

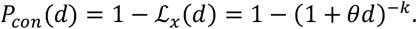

#### Gamma + point mass model

If a population has two proportion where a proportion *p*_1_ has the level *a*_1_ while the other proportion (1 − *p*_1_) follows gamma distribution:

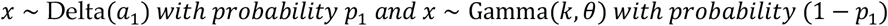

and, using its Laplace transform, the probability of infection is

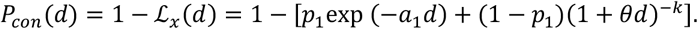

#### Gamma + point mass at zero model

If a population has two proportion where a proportion *p*_1_ is completely immune (i.e., the level 0) while the other proportion (1 − *p*_1_) follows gamma distribution:

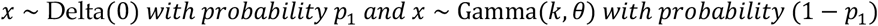

and, using its Laplace transform, the probability of infection is

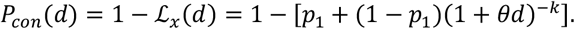

The results of model fits are summarized in **Table S1** and **S2**. For the detailed description of Tweedie model, see the main text.

## Supplementary Materials for

**Table S1**. Model comparison of dose-response models for unvaccinated individuals employing different distributions

**Table S2**. Model comparison of dose-response models for vaccinated individuals employing different distributions.

**Table S3**. Collected data on wild-type influenza human challenge studies with unvaccinated individuals

**Table S4**. Collected data on wild-type influenza human challenge studies with vaccinated individuals

## Supporting figures and tables

**Table S1.**
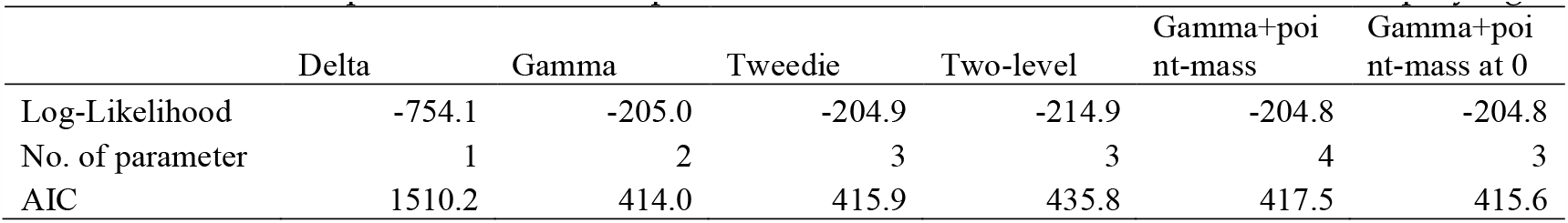
Model comparison of dose-response models for unvaccinated individuals employing different distributions.

|  | Delta | Gamma | Tweedie | Two-level | Gamma+poi<br>nt-mass | Gamma+poi<br>nt-mass at 0 |
| --- | --- | --- | --- | --- | --- | --- |
| Log-Likelihood | -754.1 | -205.0 | -204.9 | -214.9 | -204.8 | -204.8 |
| No. of parameter | 1 | 2 | 3 | 3 | 4 | 3 |
| AIC | 1510.2 | 414.0 | 415.9 | 435.8 | 417.5 | 415.6 |

**Table S2.**
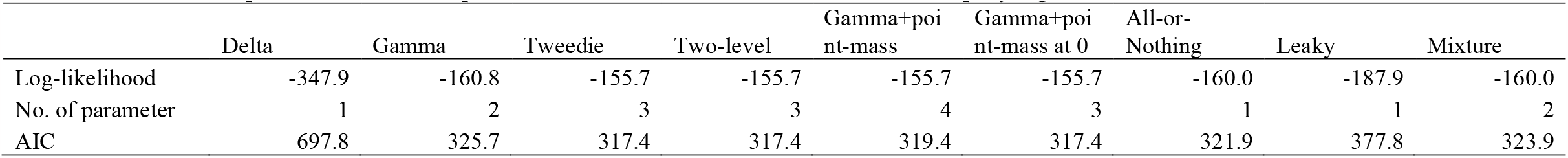
Model comparison of dose-response models for vaccinated individuals employing different distributions.

**Table S3.** Wild-type influenza human challenge studies with unvaccinated individuals.

| Dose | Total | Infected | Unit | Type/Subtype | Strain | Vaccine | Inoculation | Reference |
| --- | --- | --- | --- | --- | --- | --- | --- | --- |
| 1000000 | 24 | 23 | TCID50<sup>†</sup> | A/H3N2 | Washington/897/80 | No | Nasal | 29 |
| 1000000 | 27 | 25 | TCID50 | A/H3N2 | Washington/897/80 | No | Nasal | 30 |
| 10000 | 15 | 11 | TCID50 | A/H1N1 | California/10/78 | No | Nasal | 30 |
| 2511886 | 28 | 26 | TCID50 | A/H1N1 | Texas/1/85 | No | Nasal | 31 |
| 10000000 | 10 | 10 | TCID50 | A/H3N2 | Bethesda/1/85 | No | Nasal | 31 |
| 10000000 | 12 | 10 | TCID50 | B | Ann Arbor/1/66 | No | Nasal | 32 |
| 10000000 | 12 | 7 | TCID50 | A/H1N1 | Texas/36/91 | No | Nasal | 33 |
| 10000000 | 4 | 3 | TCID50 | A/H3N2 | Shangdong/9/93 | No | Nasal | 33 |
| 10000000 | 8 | 5 | TCID50 | B | Panama/45/90 | No | Nasal | 33 |
| 10000 | 15 | 11 | TCID50 | A/H1N1 | California/10/78 | No | Nasal | 34 |
| 31623 | 9 | 8 | TCID50 | A/H1N1 | California/10/78 | No | Nasal | 34 |
| 15849 | 6 | 6 | TCID50 | A/H1N1 | Hong Kong/123/77 | No | Nasal | 34 |
| 10000000 | 14 | 14 | TCID50 | A/H1N1 | Kawasaki/9/86 | No | Nasal | 34 |
| 10000000 | 16 | 16 | TCID50 | A/H1N1 | Kawasaki/9/86 | No | Nasal | 34 |
| 2511886 | 28 | 26 | TCID50 | A/H1N1 | Texas/1/85 | No | Nasal | 34 |
| 5011872 | 22 | 20 | TCID50 | A/H1N1 | Texas/1/85 | No | Nasal | 34 |
| 100000 | 19 | 19 | TCID50 | A/H1N1 | Texas/36/91 | No | Nasal | 34 |
| 100000 | 8 | 8 | TCID50 | A/H1N1 | Texas/36/91 | No | Nasal | 34 |
| 100000 | 8 | 8 | TCID50 | A/H1N1 | Texas/36/91 | No | Nasal | 34 |
| 1000000 | 12 | 8 | TCID50 | A/H1N1 | Texas/36/91 | No | Nasal | 34 |
| 1000000 | 16 | 13 | TCID50 | A/H1N1 | Texas/36/91 | No | Nasal | 34 |
| 1000000 | 18 | 17 | TCID50 | A/H1N1 | Texas/36/91 | No | Nasal | 34 |
| 1000000 | 19 | 14 | TCID50 | A/H1N1 | Texas/36/91 | No | Nasal | 34 |
| 100000 | 33 | 24 | TCID50 | A/H1N1 | Texas/91 | No | Nasal | 34 |
| 100000 | 26 | 25 | TCID50 | A/H1N1 | Texas/91 | No | Nasal | 34 |
| 15849 | 8 | 8 | TCID50 | A/H3N2 | Alaska/6/77 | No | Nasal | 34 |
| 10000000 | 10 | 10 | TCID50 | A/H3N2 | Bethesda/1/85 | No | Nasal | 34 |
| 14125375 | 19 | 18 | TCID50 | A/H3N2 | Bethesda/1/85 | No | Nasal | 34 |
| 25118864 | 7 | 7 | TCID50 | A/H3N2 | Bethesda/1/85 | No | Nasal | 34 |
| 1000000 | 14 | 12 | TCID50 | A/H3N2 | Korea/82 | No | Nasal | 34 |
| 10000000 | 24 | 22 | TCID50 | A/H3N2 | Los Angeles/2/87 | No | Nasal | 34 |
| 158489 | 19 | 19 | TCID50 | A/H3N2 | Victoria/3/75 | No | Nasal | 34 |
| 1000000 | 27 | 25 | TCID50 | A/H3N2 | Washington/897/80 | No | Nasal | 34 |
| 126 | 3 | 0 | TCID50 | A/H3N2 | Bethesda/10/63 | No | Aerosol | 35 |
| 78 | 3 | 0 | TCID50 | A/H3N2 | Bethesda/10/63 | No | Aerosol | 35 |
| 59 | 3 | 1 | TCID50 | A/H3N2 | Bethesda/10/63 | No | Aerosol | 35 |
| 1 | 1 | 1 | TCID50 | A/H3N2 | Bethesda/10/63 | No | Aerosol | 35 |
| 2 | 4 | 1 | TCID50 | A/H3N2 | Bethesda/10/63 | No | Aerosol | 35 |
| 5 | 9 | 4 | TCID50 | A/H3N2 | Bethesda/10/63 | No | Aerosol | 35 |

<sup>†</sup>TCID50 : Median tissue culture infectious dose.

**Table S4.** Wild-type influenza human challenge studies with vaccinated individuals.

| Dose | Total | Infected | Unit | Type/Subtype | Strain | Vaccine | Inoculation | Reference |
| --- | --- | --- | --- | --- | --- | --- | --- | --- |
| 1000000 | 16 | 3 | TCID50 | A/H3N2 | Washington/897/80 | Yes | Nasal | 29 |
| 1000000 | 16 | 10 | TCID50 | A/H3N2 | Washington/897/80 | Yes | Nasal | 29 |
| 1000000 | 16 | 11 | TCID50 | A/H3N2 | Washington/897/80 | Yes | Nasal | 30 |
| 1000000 | 16 | 11 | TCID50 | A/H3N2 | Washington/897/80 | Yes | Nasal | 30 |
| 10000 | 14 | 7 | TCID50 | A/H1N1 | California/10/78 | Yes | Nasal | 30 |
| 10000 | 18 | 12 | TCID50 | A/H1N1 | California/10/78 | Yes | Nasal | 30 |
| 2511886 | 20 | 12 | TCID50 | A/H1N1 | Texas/1/85 | Yes | Nasal | 31 |
| 2511886 | 16 | 7 | TCID50 | A/H1N1 | Texas/1/85 | Yes | Nasal | 31 |
| 10000000 | 11 | 5 | TCID50 | A/H3N2 | Bethesda/1/85 | Yes | Nasal | 31 |
| 10000000 | 13 | 9 | TCID50 | B | Ann Arbor/1/66 | Yes | Nasal | 32 |
| 10000000 | 10 | 3 | TCID50 | A/H1N1 | Texas/36/91 | Yes | Nasal | 33 |
| 10000000 | 10 | 2 | TCID50 | A/H1N1 | Texas/36/91 | Yes | Nasal | 33 |
| 10000000 | 8 | 4 | TCID50 | A/H3N2 | Shangdong/9/93 | Yes | Nasal | 33 |
| 10000000 | 10 | 3 | TCID50 | A/H3N2 | Shangdong/9/93 | Yes | Nasal | 33 |
| 10000000 | 7 | 2 | TCID50 | B | Panama/45/90 | Yes | Nasal | 33 |
| 10000000 | 7 | 0 | TCID50 | B | Panama/45/90 | Yes | Nasal | 33 |
| 127 | 15 | 4 | TCID50 | A/H3N2 | Aichi/2/68 | Yes | Nasal | 36 |
| 127 | 14 | 0 | TCID50 | A/H3N2 | Aichi/2/68 | Yes | Nasal | 36 |

†TCID50 : Median tissue culture infectious dose.

